# Placental pathology in COVID-19

**DOI:** 10.1101/2020.05.08.20093229

**Authors:** Elisheva D. Shanes, Leena B. Mithal, Sebastian Otero, Hooman A. Azad, Emily S. Miller, Jeffery A. Goldstein

## Abstract

**Objectives:** To describe histopathologic findings in the placentas of women with COVID-19 during pregnancy.

**Methods:** Pregnant women with COVID-19 delivering between March 18, 2020 and May 5, 2020 were identified. Placentas were examined and compared to historical controls and women with placental evaluation for a history of melanoma.

**Results:** 16 placentas from patients with SARS-CoV-2 were examined (15 with live birth in the 3^rd^ trimester 1 delivered in the 2^nd^ trimester after intrauterine fetal demise). Compared to controls, third trimester placentas were significantly more likely to show at least one feature of maternal vascular malperfusion (MVM), including abnormal or injured maternal vessels, as well as delayed villous maturation, chorangiosis, and intervillous thrombi. Rates of acute and chronic inflammation were not increased.

The placenta from the patient with intrauterine fetal demise showed villous edema and a retroplacental hematoma.

**Conclusions:** Relative to controls, COVID-19 placentas show increased prevalence of features of maternal vascular malperfusion (MVM), a pattern of placental injury reflecting abnormalities in oxygenation within the intervillous space associated with adverse perinatal outcomes. Only 1 COVID-19 patient was hypertensive despite the association of MVM with hypertensive disorders and preeclampsia. These changes may reflect a systemic inflammatory or hypercoagulable state influencing placental physiology.

**Key Points:**

1. The placentas of women infected with SARS-CoV2 have higher rates maternal vascular malperfusion features compared to controls.
2. Maternal vascular malperfusion has been associated with adverse perinatal outcomes, such as preeclampsia, fetal growth restriction, preterm birth, and stillbirth.
3. As the placentas of women with SARS-CoV2 show reproducible histopathologic abnormalities, these findings suggest increased antenatal surveillance for women with COVID-19 may be warranted.

## Introduction

The impact of the severe acute respiratory syndrome coronavirus 2 (SARS-CoV-2) and the associated disease, COVID-19, on pregnant women and infants is of particular interest to obstetricians, pediatricians, and patients.

Coronaviruses are single-stranded, encapsulated RNA viruses. Most of the commonly circulating coronaviruses cause mild upper respiratory infection with occasional severe illness in immunocompromised individuals. Previous novel coronaviruses causing significant human infection include the severe acute respiratory syndrome (SARS, caused by the coronavirus SARS-CoV) from 2002-2004 and Middle East respiratory syndrome (MERS, caused by the coronavirus MERS-CoV) from 2012 to present. The SARS epidemic (officially ending in 2003, though sporadic cases were reported into 2004) is estimated to have involved approximately 100 pregnant women worldwide.^1^ Several case series demonstrated that SARS infection during pregnancy was associated with severe maternal infection, increased risk of maternal death, and spontaneous abortion.^2–8^ MERS, a primarily zoonotic infection which is transmitted between humans only via very close contact, has only been confirmed in approximately 1,845 people since 2012. A case series of 5 women with MERS during pregnancy as well as subsequent case reports suggest that MERS is also associated with poor maternal and perinatal outcomes.^2,3,9^

Current case series have described clinical features and outcomes in pregnant women with COVID-19 in Wuhan, China, the original epicenter of infection.^2,6,7^ Taken together, the body of evidence does not currently suggest that pregnant women are subject to more severe disease, in contrast to both SARS and MERS. However adverse perinatal outcomes have been reported, including increased risks of miscarriage, preeclampsia, preterm birth, and stillbirth.^4^

Histopathologic examination of placental tissue can contribute significant information regarding the health of both mother and fetus. A variety of viral infections in pregnancy are associated with specific placental findings, including lymphoplasmacytic villitis with associated enlargement of villi and intravillous hemosiderin deposition in the setting of maternal cytomegalovirus (CMV) infection^10^, as well as rare reports of intervillositis in the setting of Zika virus^11,12^ and Dengue virus^13^, among others.

While there are no known placental findings associated with the common coronaviruses, Ng et al. reported placental pathology in seven women with SARS infection in Hong Kong.^14^ The placentas of three women who delivered while acutely ill demonstrated increased perivillous or subchorionic fibrin, while in two women who had recovered from third trimester infection by the time of delivery, there were large zones of avascular villi, with one of the two additionally demonstrating a large villous infarct; both contained increased nucleated red blood cells in the fetal circulation. None of the seven placentas examined had any acute or chronic inflammatory processes. While there are no histopathologic studies of placental findings in MERS, there is one report^15^ describing placental abruption in this setting.

The current COVID-19 pandemic is still in its early stages, with preliminary case series of infection in pregnant women beginning to become available. The Chinese literature includes one report of placental findings in three women with COVID-19.^16^ The authors describe increased perivillous fibrin deposition in all three placentas, multiple villous infarcts in one placenta, and a chorangioma in another case. While there is one case report of second trimester miscarriage in a mother with COVID in which placental findings (acute inflammation and increased perivillous fibrin),^17^ there are, to the best of our knowledge, no published case series in the English literature of placental pathology in women diagnosed with COVID-19 during pregnancy. In this report, we present placental findings in 16 women with SARS-CoV-2 infection during pregnancy and compare results to historical controls.

## Materials and Methods

### Patients

Pregnant women with COVID-19 delivering between March 18, 2020 and May 5, 2020 were identified via the electronic health record and tracked using REDCap.^18^ From March 18, 2020 until April 7, 2020, only women with moderate to severe symptoms of COVID-19 underwent testing. After April 7, 2020, all women presenting to labor and delivery were universally tested. All patients were tested using nasopharyngeal swab placed in M4 viral transport medium. Prior to April 7, 2020, testing was performed using a laboratory-developed version of the Centers for Disease Control and Prevention (CDC) SARS-CoV-2 reverse transcriptase polymerase chain reaction (RT-PCR) assay.^19^ Total RNA was extracted by Qiagen MinElute Virus Spin Kit. Reverse transcription was performed by TaqPath 1 StepRT-qPCR MM CG (A15299) using the 2019-nCOV Kit (IDT 10006606). PCR was performed on Quant Studio 6 flex. The reported limit of detection is 1000 copies / ml. After April 7, 2020, patients were tested using the GeneXpert Dx Xpress SARS-CoV-2 RT-PCR assay (Cepheid, Sunnyvale, CA). The analytical sensitivity and specificity are reported by the manufacturer as 100% (87/87 samples) and 100% (30/30 samples) with a limit of detection of 250 copies /ml or 0.0100 plaque-forming units per ml.^20^ Positive SARS-CoV-2 test was considered an independent criteria for submission of placentas to pathology.

### Patients - Controls

Historical controls underwent singleton third trimester delivery with clinically-indicated placental pathology examination between January 1, 2011 and June 30, 2018. Examination was at the judgement of the delivering physician. Suggested criteria for examination included: Abnormal placenta and/or cord on examination in the delivery room; Stillbirth; Intrauterine growth restriction (< 10%ile); Preterm delivery ≤ 34 weeks gestation; Premature rupture of membranes < 34 weeks gestation; Severe preeclampsia; Clinical concern for viral infection; Clinical suspicion of chorioamnionitis; Clinical suspicion of abruption; History of current / previous gestational trophoblastic disease; Compromised clinical condition of neonate, including intensive care admission, cord pH <7.0, Apgar score ≤ 5 at 5 minutes, ventilatory assistance, hematocrit < 25%, hydrops fetalis, congenital anomalies, monochorionic twin gestation, all higher order multiple gestations (e.g. triplets); Maternal cancer history (including melanoma); Retained placenta. Pathology reports were retrieved from the laboratory information system. Patients with a history of melanoma have been suggested as superior controls, as this indication is orthogonal to other pregnancy complications.^19^ A nested cohort of patients with a history of melanoma was created from the historical controls based on the presence of the word “melanoma” in the clinical history section of their pathology report. Placental examination is indicated in all patients with a history of melanoma to examine for placental metastases. No patient in the study period had such metastases, so they may be considered a random sample of the delivering population. This study was approved by our Institutional Review Board, STU00207907 (historical and melanoma controls), STU00212232 (COVID-19 cases).

### Examination

Placentas underwent routine clinical examination consisting of storage at 4o C prior to fixation, fixation in 10% buffered formalin, photographs of the maternal and fetal surface, measurement, trimmed weight, sectioning and examination of the cut surface. Sections submitted included 2 of membrane rolls, at least 2 of umbilical cord, 3 maternal surface biopsies, 2 full thickness sections, and representative sampling of any lesions present. Sections underwent routine processing, embedding, sectioning at 5 μm and staining with hematoxylin and eosin. Histologic examination was performed by subspecialty perinatal pathologists who were aware of the COVID-19 status. Cases were reviewed by two pathologists to confirm the diagnoses.

### Python/NLTK/Regular expression pipeline

Our hospital’s laboratory information system (Cerner Build List Id: 2014.08.1.36) was searched for all 2nd and 3rd trimester placentas between July 2011 and June 2018. Maternal age, gestational age, and delivery mode were extracted from the clinical history within the pathology request. Final diagnoses are listed as a series of relatively self-contained bullet points, which were searched for key phrases to identify cases. To do this, individual bullet points were tokenized and stemmed using the Porter stemmer function of the Natural Language Toolkit (nltk) package (version 3.3) on Python (version 3.6.5). Stems characteristic of specific diagnoses were tested, e.g., “fibrinoid necros” for Fibrinoid Necrosis. A case was identified when all stems were present, and word order was ignored. Manual review was performed of 20-60 bullet points to test the accuracy of case identification. Over 65 diagnoses were tested for with overall sensitivity, specificity, and accuracy of 0.97, 0.97, and 0.98, respectively.

### Analysis

Comparisons were made between placentas of women with COVID-19 and historical controls as well as between placentas of women with COVID-19 and women with placental evaluation for a history of melanoma. The placenta from one second trimester miscarriage was excluded from statistical analysis. Quantities are reported as mean +/- standard deviation. Categorical data are reported as actual values or percentages. Comparisons of categorical data with patients with history of melanoma use Fisher’s exact test. Comparisons of categorical data with the larger control set use Chi squared with Yates’ correction. P-values are not corrected for multiple comparisons.

## Results

Sixteen placentas from patients with SARS-CoV-2 were examined (*Table 1*). Four patients had COVID-19 remote from delivery (i.e., infection diagnosed between 25-34 days prior to delivery), one was diagnosed 8 days prior to delivery, and the remaining 11 were diagnosed on presentation to labor and delivery. Fourteen patients delivered at term (37-40 weeks), one delivered at 34 weeks, and one represented a 16-week intrauterine fetal demise (IUFD). Five placentas were small for gestational age, one significantly (298 g, expected 409 - 589 g). One placenta was slightly large for gestational age (612 g, expected 426 – 611 g). Indications for pathologic evaluation included only positive SARS-CoV-2 in 13/16 cases; one case additionally had a history of cholestasis of pregnancy and gestational diabetes, another had a history of pregnancy-induced hypertension, and one was the aforementioned IUFD. All infants were negative for SARS-CoV-2 by nasopharyngeal and throat swab.

**Table 1:** Clinical histories of cases

| Feature | 1 | 2 | 3 | 4 | 5 | 6 | 7 | 8 | 9 | 10 | 11 | 12 | 13 | 14 | 15 | 16 |
| --- | --- | --- | --- | --- | --- | --- | --- | --- | --- | --- | --- | --- | --- | --- | --- | --- |
| **Gestational age (weeks)** | 38 | 39 | 38 | 39 | 40 | 39 | 40 | 37 | 40 | 39 | 38 | 37 | 39 | 37 | 34 | 16 |
| **Maternal age (years)** | 26 | 41 | 33 | 39 | 34 | 31 | 31 | 33 | 28 | 30 | 30 | 37 | 23 | 34 | 30 | 26 |
| **Indication for exam** | Co V | Co V | Co V | Co V | CoV | CoV | Co V | CoV | CoV | CoV | Co V | CoV Chol GDM | CoV, Asthma | Co V PIH | CoV, Asthma | IUFD, CoV PPROM |
| **Diagnosis to delivery interval (days)** | 1 | 28 | 0 | 0 | 2 | 0 | 25 | 7 | 0 | 25 | 1 | 34 | 0 | 0 | 8 | 0 |
| **Placenta weight (g)** | 298 | 612 | 357 | 463 | 400 | 424 | 557 | 542 | Frag | 606 | 544 | 519 | 394 | 423 | 416 | Frag |
| **Small / Large for gestational age** | SG A | LG A | SG A |  | SGA | SGA |  |  | N/A |  |  |  | SGA |  |  | N/A |
| **Infant SARS-CoV-2 test result** | - | - | - | - | - | - | - | - | - | - | - | - | - | - | - | - |

**Abbreviations:** Chol: cholestasis of pregnancy; CoV: SARS-CoV-2 infection; Frag: Fragmented placenta, weight unreliable; GDM: Gestational diabetes mellitus; IUFD: Intrauterine fetal demise; LGA: large for gestational age; PIH: Pregnancy induced hypertension; PPROM: Preterm premature rupture of the membranes; SGA: Small for gestational age

The 16-week IUFD occurred in an asymptomatic woman who tested positive for SARS-CoV-2 at the time of admission to labor and delivery. Placental pathology in this case demonstrated retroplacental hematoma, and villous edema; villous maturation was appropriate for gestational age. There was no acute or chronic inflammation noted. This case is not included in the cohort for statistical analysis as the control population is of third trimester deliveries only.

Fifteen cases were compared with the two control populations described previously. Features of maternal vascular malperfusion (MVM) were present in 12/15 cases, significantly higher than melanoma (59/215, p = 0.001, *Table 2, Figure*) as well as all controls (7754/17479, p = 0.046). Features included central (1/15) and peripheral (3/15) villous infarctions, villous agglutination (3/15), atherosis and fibrinoid necrosis of maternal vessels (3/15), mural hypertrophy of membrane arterioles (5/15), and accelerated villous maturation (2/15). Peripheral infarctions, atherosis and fibrinoid necrosis, and mural hypertrophy were more common in COVID-19 cases than in placentas of women with a history of melanoma (p = 0.01, p = 0.002, p = 0.01, respectively), while atherosis and fibrinoid necrosis and mural hypertrophy were more common in COVID-19 cases than in all historical controls (p = 0.001 and p = 0.001, respectively).

**Figure 1:**
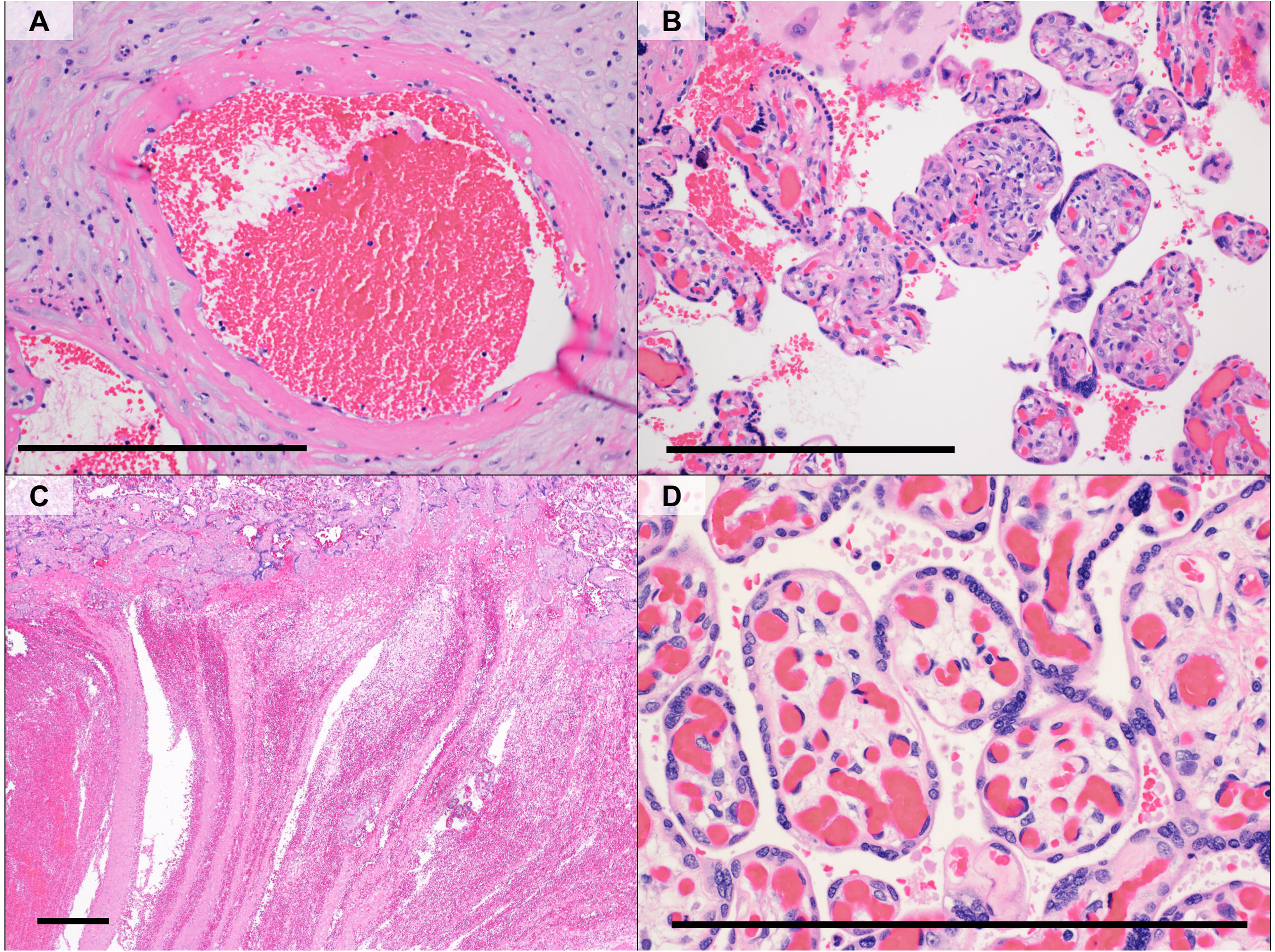
Selected pathology from COVID-19 patients. A) Maternal arteriole with atherosis and fibrinoid necrosis; B. Small focus of fetal villi with lymphocytic villitis; C) Intervillous thrombus showing lines of Zahn; D) Fetal villi with chorangiosis and edema. Scale bar 0.5 mm. Original magnifications 20x, 20x, 5x, and 40x.

**Table 2:**
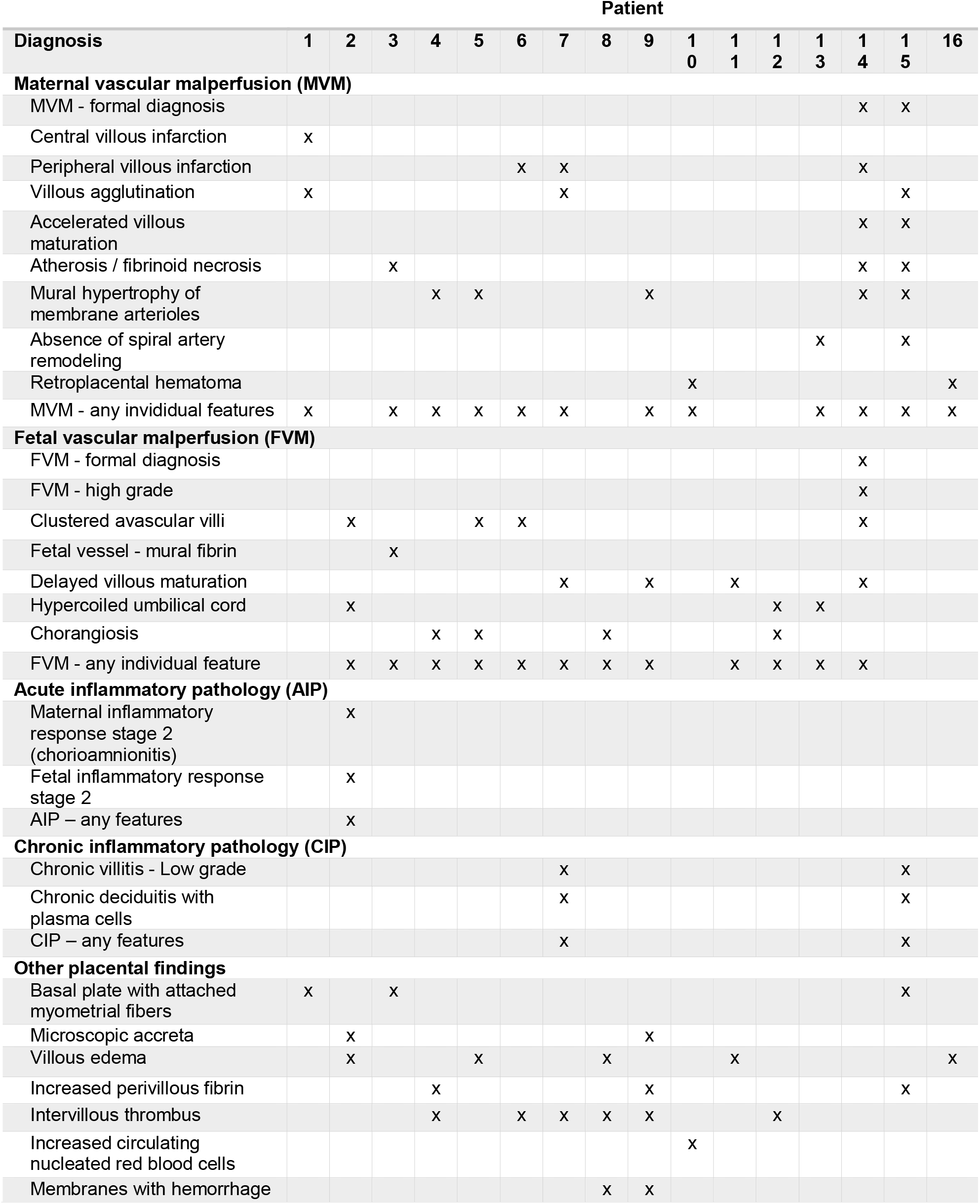
Diagnoses

**Table 3:**
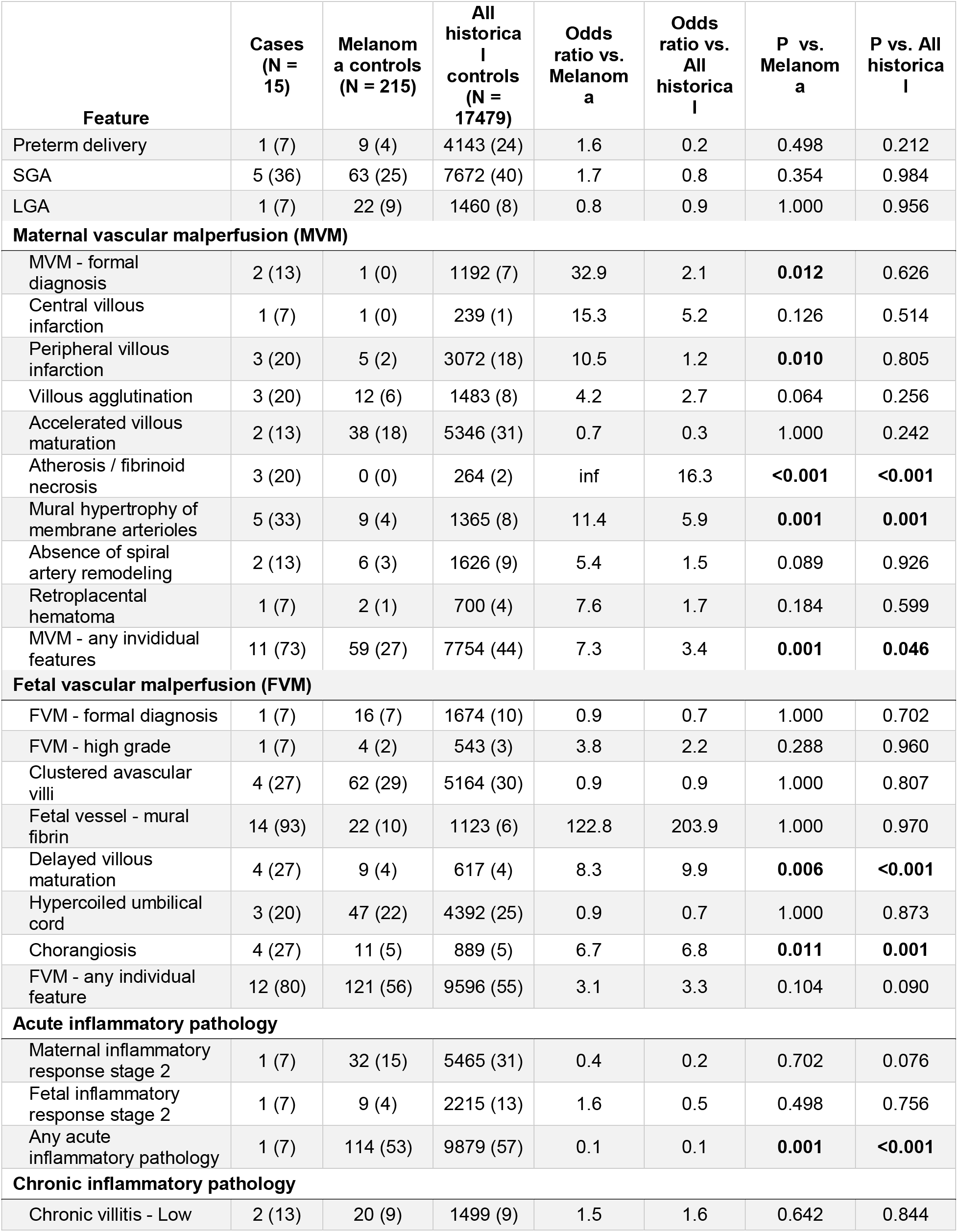
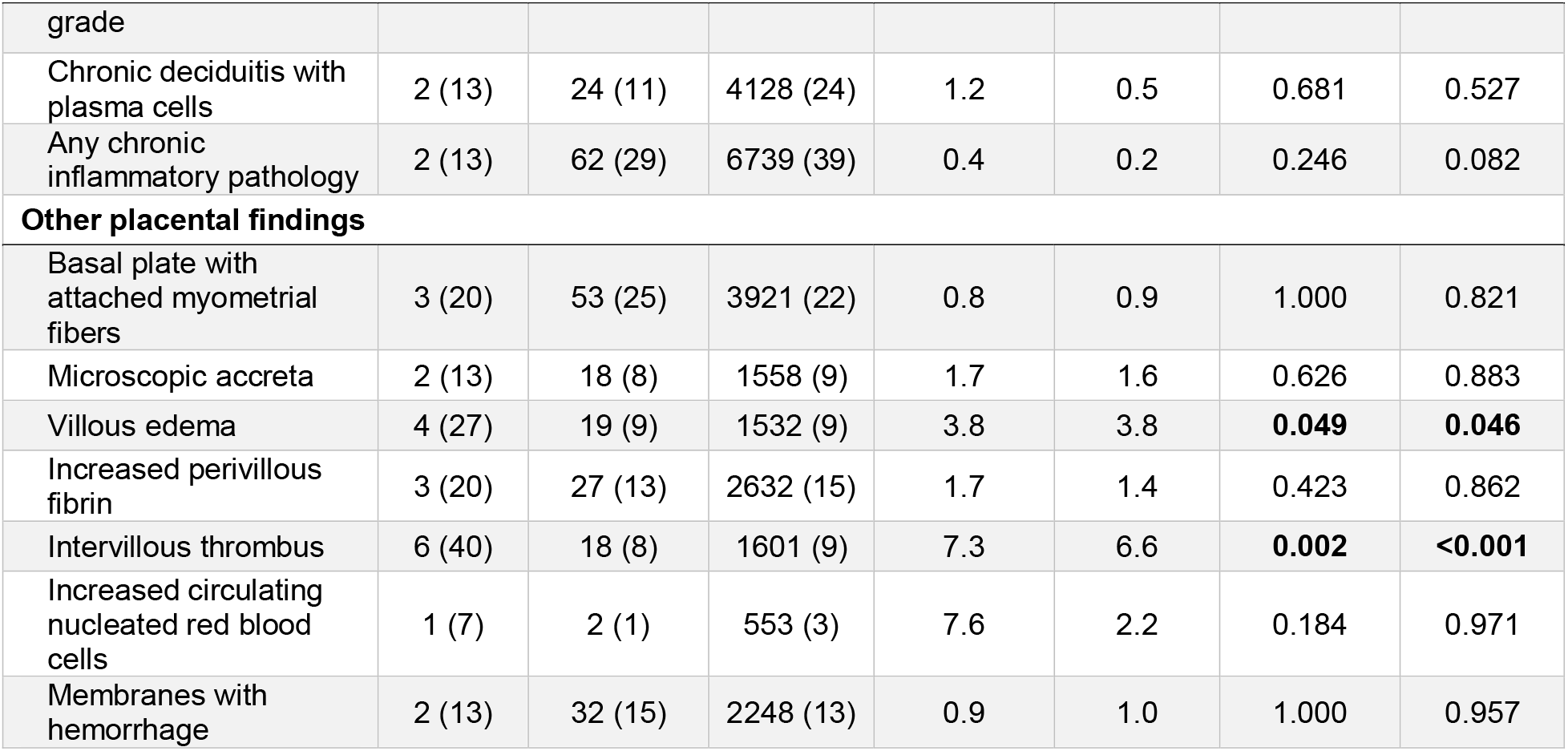
Comparison of SARS-CoV-2 patients with controls

Features of fetal vascular malperfusion (FVM) were present in 12/15 cases, not significantly different from melanoma (121/215, p = 0.10) or all historical (9596 / 17479, p = 0.09) controls. Specifically, 4/15 COVID-19 placentas showed clustered avascular villi, 1/15 showed mural fibrin deposition in fetal vessels, 4/15 showed delayed villous maturation, and 3/15 showed a hypercoiled umbilical cord. 4/15 placentas showed chorangiosis, which was increased relative to all historical controls (889/17479, p = 0.001), as well as melanoma controls (11/215, p = 0.011).

One COVID-19 case showed acute inflammatory pathology (AIP) including histologic chorioamnionitis and umbilical arteritis, corresponding to maternal and fetal inflammatory response stages of 2 and 2, respectively. The rate of acute inflammatory pathology is significantly lower than both melanoma (114/215, p = 0.001) and all historical (9879/17479, p = 0.0003) controls.

Two COVID-19 cases showed chronic inflammatory pathology (CIP), both with low grade chronic lymphocytic villitis and chronic deciduitis with plasma cells. These rates are not significantly different from melanoma (62/215, p = 0.246) or all historical (6739/17479, p =0.082) controls.

Additional findings in the COVID-19 cases included 3 cases of basal plate with adherent myometrial fibers, 2 cases of microscopic accreta, 3 with increased perivillous fibrin deposition, and 1 with increased circulating nucleated red blood cells. 5 cases demonstrated villous edema, significantly increased from both melanoma (12/215, p = 0.049) and all historical (1532/17479, p = 0.046) controls. 6/15 COVID-19 cases contained intervillous thrombi, significantly increased relative to melanoma (18/215, p = 0.002) and all historical (1601/17479, p = 0.0002) controls.

## Discussion

This is the first case series in the English language literature of placental pathology in SARS-CoV-2 infection and COVID-19 of which we are aware. Despite the relatively small number of cases, some trends emerge. MVM, previously known as maternal vascular underperfusion, has been associated with oligohydramnios, fetal growth restriction, preterm birth, and stillbirth.^20–23^ Maternal hypertensive disorders, including gestational hypertension and preeclampsia, are the major risk factors for MVM.^24,25^

Surprisingly, only one of the patients in our study was hypertensive. This suggests a relationship between SARS-CoV-2 infection and MVM, though the causality is difficult to parse. The histologic changes of MVM are thought to represent some chronicity, though exact timing is unknown, and these features can be seen in women who develop preeclampsia only during or after childbirth. Whether systemic vascular changes due to maternal COVID-19 are responsible for the histologic changes of MVM cannot be determined.

As SARS-CoV-2 is a virus, it might be expected to induce inflammation. CIP and in particular chronic villitis may be caused directly by some viral infections, such as cytomegalovirus.^10^ Chronic villitis also shows seasonality, suggesting a link to circulating viruses, either causing direct infection or loss of tolerance.^26^ In that context, it is relevant that neither AIP nor CIP were increased in COVID-19 patients relative to the controls. Indeed, both categories of disease were less prevalent in COVID-19 patients, AIP significantly so. None of the COVID-19 patients in our study were severely ill or undergoing a “cytokine storm” and it may be possible that CIP could be induced in those cases of systemic inflammation.

COVID-19 patients showed a significant increase in intervillous thrombi. The intervillous space usually contains flowing maternal blood, yet 85% of thrombi are suggested to be of fetal origin.^27^ Intervillous thrombi are generally considered incidental findings, but have been associated with maternal hypertensive disorders or co-incident infarctions.^28–30^ In the context of research suggesting an increase of thrombotic and thromboembolic disorders in COVID-19, these may represent placental formation or deposition of thrombi in response to the virus.^31–33^

The increased incidence of chorangiosis is notable as well. Chorangiosis is associated with decreased maternal oxygen saturation; ‘ it is more commonly seen in women living at high altitudes, as well as, for unknown reasons, in women with diabetes.^37^ Two of the cases with chorangiosis in the current study represent asymptomatic women, while one is from a woman with one week of symptomatic COVID-19, and the fourth is from a woman who recovered from symptomatic COVID-19 diagnosed initially 34 days prior to delivery. The latter woman also had gestational diabetes, complicating interpretation of the findings. While the association between a respiratory virus and a finding sometimes seen in maternal hypoxia is interesting, the small sample size and confounding factors make it difficult to draw any conclusions at this time.

Comparison with the reported features of three placentas in the Chinese-language literature are of interest.^16^ In the previous study, the three patients had variably increased perivillous fibrin deposition in comparison to only one in our series. We also report villous infarction in several placentas, reinforcing the significance of MVM lesions. Finally, the previous study reported one case of chorangioma, which may be incidental but has been associated with both FVM and MVM changes,^38,39^ and may have a relationship to chorangiosis, which was significantly increased in the present study.

Our cohort includes a 16-week IUFD, which was excluded from the subsequent statistical analysis. Placental pathology demonstrated villous edema and a retroplacental hematoma. Retroplacental hematomas are a feature of MVM, however in this case it may have been procedural.^21^ In comparison, a second trimester miscarriage has been reported in a patient with COVID-19.^17^ The placenta in this case showed mildly increased perivillous fibrin and acute subchorionitis. SARS-CoV-2 nucleic acid testing was negative in the fetus but positive in placenta. Bacterial culture and PCR were negative, leading the authors to suggest the acute inflammatory response is related to SARS-CoV-2. We do not see an increased incidence of acute inflammatory pathology in our IUFD or overall in our cases. Low stage inflammation is common in laboring deliveries and investigation for bacteria is frequently negative.^40,41^ A larger case series is needed to define the relationship between SARS-CoV-2, acute inflammation and IUFD.

None of the placentas in our study were directly tested for SARS-CoV-2 viral RNA. Per hospital protocol, all infants born to SARS-CoV-2 mothers were tested by a nasopharyngeal and throat swab at > 24 hours of life a minimum of once and up to twice for SARS-CoV-2, and none were positive. This corroborates existing evidence that vertical transmission of the virus is uncommon, and suggests that placental changes, if caused by COVID-19, are related to maternal infection and inflammation rather than fetal infection.

In determining which placental anomalies are attributable to SARS-CoV-2 infection, we would argue that statistically significant associations identified in controlled studies, such as this one, are the appropriate first step. The scope of placental anomalies is vast, and many anomalies are frequently seen across a variety of indications for placental examination, emphasizing the need for appropriate controls.^38,42^

This study is subject to some limitations. The relatively low number of patients limits our assessment of low frequency or variable outcomes. We have grouped symptomatic, asymptomatic, and recovered patients - a larger study could treat these groups separately. One strength of our study is the use of 2 distinct control groups. Placentas are only submitted for clinical examination when there is an indication - usually diseases in pregnancy or complications of labor and delivery. Melanoma history is an indication for placental examination but is considered independent of pregnancy outcome, and therefore some have advocated using these patients as a control. Nonetheless, more appropriate control populations could include patients with influenza-like illness with negative SARS-CoV-2 testing and asymptomatic, COVID-19 negative patients delivering at a similar time. Finally, our study does not formally test causality or the direct relationship between SARS-CoV-2 infection and development of placental pathology. It is possible that viral infection directly leads to placental pathology or that there is a common underlying cause for both placental lesions and susceptibility to SARS-CoV-2.

## Conclusions

We report placental pathology from 16 patients with SARS-CoV-2 infection. No pathognomonic features are identified, however there are increased rates of maternal vascular malperfusion features and intervillous thrombi, suggesting a common theme of abnormal maternal circulation, as well as an increased incidence of chorangiosis. These findings provide mechanistic insight into the observed epidemiologic associations between COVID-19 in pregnancy and adverse perinatal outcomes. Collectively, these findings suggest that increased antenatal surveillance for women diagnosed with SARS-CoV-2 may be warranted.

## Data Availability

Original data are available following execution of a Data Use Agreement with Northwestern University.

## Acknowledgements

The authors wish to thank Dr. Kruti Maniar (Northwestern Department of Pathology), staff in the Obstetric-COVID unit at Prentice Women’s Hospital and the Northwestern Memorial Hospital Department of Pathology.

